# A unique cytotoxic CD4^+^ T cells signature defines critical COVID-19

**DOI:** 10.1101/2023.02.17.23286059

**Authors:** Sarah Baird, Caroline L. Ashley, Felix Marsh-Wakefield, Sibel Alca, Thomas M. Ashhurst, Angela L. Ferguson, Hannah Lukeman, Claudio Counoupas, Jeffrey J. Post, Pamela Konecny, Adam Bartlett, Marianne Martinello, Rowena A. Bull, Andrew Lloyd, Alice Grey, Owen Hutchings, Umaimainthan Palendira, Warwick J. Britton, Megan Steain, James A. Triccas

**Affiliations:** School of Medical Sciences, Faculty of Medicine and Health, The University of Sydney, Camperdown, NSW, Australia; Sydney Institute for Infectious Diseases and the Charles Perkins Centre, The University of Sydney, Camperdown, NSW, Australia; Vascular Immunology Unit, The University of Sydney, Sydney, New South Wales, Australia; Sydney Cytometry Core Research Facility, Charles Perkins Centre, Centenary Institute and The University of Sydney; Tuberculosis Research Program, Centenary Institute, Sydney, NSW, Australia; Prince of Wales Clinical School, UNSW Australia, Sydney, NSW Australia; St George Hospital, Sydney, New South Wales, Australia; The Kirby Institute, UNSW, Sydney, NSW, Australia; School of Biomedical Sciences, Faculty of Medicine, UNSW, Sydney, NSW, Australia; Sydney Children’s Hospital, Sydney, NSW, Australia; RPA Virtual Hospital, Sydney Local Health District, Sydney, NSW, Australia; Department of Clinical Immunology, Royal Prince Alfred Hospital, Camperdown, Australia

**Keywords:** SARS-CoV-2, COVID-19, T cells, Spectral Cytometry, CD4-CTLs

## Abstract

**Background and objectives:** SARS-CoV-2 infection causes a spectrum of clinical disease presentation, ranging from asymptomatic to fatal. While neutralising antibody (NAb) responses correlate with protection against symptomatic and severe infection, the contribution of the T cell response to the resolution or progression of disease is still unclear. Optimal protective immunity may require activation of distinct immune pathways. As such, defining the contribution of individual T cell subsets to disease outcome is imperative to inform the development of next-generation COVID-19 vaccines. To address this, we performed immunophenotyping of T cell responses in unvaccinated individuals, representing the full spectrum of COVID-19 clinical presentation.

**Methods:** Spectral cytometry was performed on peripheral blood mononuclear cell samples from patients with PCR-confirmed SARS-CoV-2 infection. Computational and manual analyses were used to identify T cell populations associated with distinct disease states through unbiased clustering, principal component analysis and discriminant analysis.

**Results:** Critical SARS-CoV-2 infection was characterised by an increase in activated and cytotoxic CD4^+^ (CTL) cells of a T follicular helper (T_FH_) or effector memory re-expressing CD45RA (T_EMRA_) phenotype. These CD4^+^ CTLs were largely absent in those with less severe disease. In contrast, those with asymptomatic or mild disease were associated with high proportions of naïve T cells and reduced expression of activation markers.

**Conclusion:** Highly activated and cytotoxic CD4^+^ T cell responses may contribute to cell-mediated host tissue damage and progression of COVID-19. Potential for induction of these detrimental T cell responses should be considered when developing and implementing effective COVID-19 control strategies.

## Introduction

The Severe acute respiratory syndrome coronavirus 2 (SARS-CoV-2) pandemic has been ongoing since March of 2020. As of February 2023, over 754 million cases of SARS-CoV-2 infection and 6.83 million fatalities from coronavirus disease 2019 (COVID-19) have been reported.^1^ While several vaccines are now available for use, SARS-CoV-2 remains a leading cause of infectious disease death globally. One of the major challenges with SARS-CoV-2 infection is the spectrum of COVID-19 clinical presentation, ranging from asymptomatic to fatal. It is thought that more severe disease results from a dysregulated immune response to infection; however, variability in this immune dysfunction between individuals has limited understanding of the correlates of disease severity. Developing a more comprehensive understanding of the immune response across the spectrum of COVID-19 clinical presentation will help to differentiate protective from pathogenic immune responses. This is essential to inform the development of next-generation therapies and vaccines against SARS-CoV-2, with improved longevity and efficacy against newly emerging variants of concern (VOC).

The key correlate of protective immunity against infection and severe disease in COVID-19 is neutralising antibody responses (NAb).^2,3^As such, the factors that contribute to breakthrough infection following vaccination centre around humoral immune responses, such as waning NAb titres, and antibody escape mutations on globally dominant VOC.^4–9^ The T cell response appears to have greater longevity than detectable NAbs, with sustained response to antigen stimulation demonstrated >1-year post-infection.^10,11^ Additionally, the dominant T cell epitopes do not overlap with areas of high mutation on variant viruses, and as a result the T cell response is preserved against antibody-escape VOC. ^9,12–16^ Considering the limitations of current vaccines, it has been suggested that the long-lived T cell response against SARS-CoV-2 variants may contribute to protective immunity in the absence of a robust humoral immune response.^12,13,16^ While NAb responses have been shown to tightly correlate with protection against disease, no such correlation has been shown with the T cell response to SARS-CoV-2.^17^ There is evidence that polyfunctional and cross-reactive T cell responses to seasonal coronaviruses are associated with milder disease and faster viral clearance.^18–20^ However, several studies have also described an expansion of highly activated T cells in severe COVID-19, that could potentially contribute to excessive inflammatory immune responses and host-tissue damage.^21–23^ As such, whether T cells play a protective or pathogenic role in COVID-19 is still unresolved.

To better define the role of T cell subsets, we performed an explorative investigation into T cell phenotypes across the clinical spectrum of COVID-19 presentation, utilising an unbiased analysis approach with a T-cell-centric high-dimensional cytometry panel. We report that critical COVID-19 infection is characterised by a shift from naïve T cell phenotypes to an expansion of cytotoxic CD4^+^ T lymphocyte subsets.

## Results

### The T cell compartment distinguishes critical SARS-CoV-2 infection from other disease states

To obtain a global view of the T cell response within and between disease states, peripheral blood mononuclear cells (PBMCs) were isolated from the blood of patients and spectral cytometry was performed using a T cell-centric antibody panel. Initially, T cell populations were manually gated (Figure s1) and differences in the proportion of each population between patients were identified by an unsupervised Principal Component Analysis (PCA), where each data point represents one patient sample (Figure 1a). When the COVID-19 severity of each patient was superimposed onto the PCA patients with critical infection separated distinctly across the first component (dim 1, accounted for 31.8% of the variance) from most other patient samples (Figure 1a). Visualisation of the contribution of each T cell population proportion to the principal components revealed that the expression of activation/proliferation markers HLA-DR, Granzyme B (GZMB), Perforin (PFN), and Ki-67 on central memory (T_CM_), effector memory (T_EM_), and effector memory re-expressing CD45RA (T_EMRA_) contribute to the separation of samples in dimension 1 (Figure 1b). Furthermore, the proportions of these activated, memory CD4^+^ and CD8^+^ T cell subsets were negatively correlated to those of CD4^+^ and CD8^+^ naïve T cells (T_N_) (Figure 1b).

**Figure 1.**
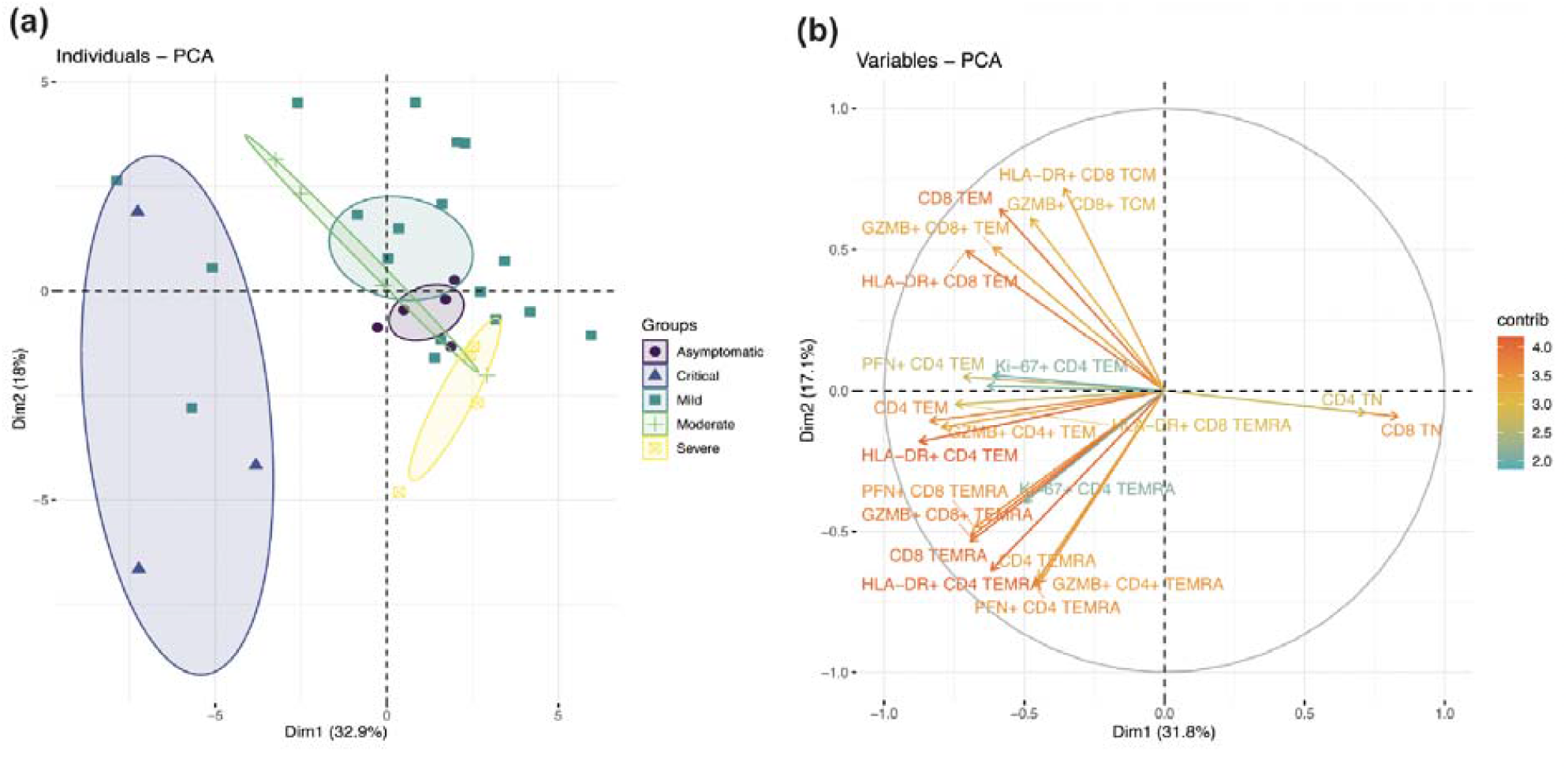
Variance in the T cell compartment explained by COVID-19 severity. **A)** Principal component analysis based on the relative abundance of 38 T cell populations in patients with asymptomatic to critical COVID-19 (n = 35). Patient disease severity overlay on principal component analysis where non-overlapping disease states represent a difference in the relative abundance of T cell populations between groups. **B)** Variable contribution plot visualising the T cell populations that contribute to the principal components. Arrow direction represents correlation, where opposing direction is negative and adjacent arrows represent positive correlation between variables. Differences between groups determined by FDR.

### Metaclusters of cytotoxic CD4^+^ T cells differentiate critical and severe SARS-CoV-2 infection

To determine which T cell populations may be implicated in progression from severe to critical COVID-19, patients in those groups were selected for further analysis. To fully capture the heterogeneity of activation and cytotoxic marker expression in the T cell compartment of these patients, unbiased clustering was performed. FlowSOM clustering was set to create 25 metaclusters (Mcs) and fast interpolation-based t-SNE (Fit-SNE) was used to visualise the proportion of each Mc (Figure 2a & b), with a heatmap generated for phenotyping (Figure 2c). As the T-cell panel included markers to define conventional CD4^+^ and CD8^+^ subsets, CD4^-^CD8^-^ Mcs (7, 8, 11, 14 and 22) were excluded and a Partial Least Squares Discriminant Analysis (PLS-DA) was performed to identify the Mcs that contribute to variance between severe and critical infection. Like the PCA (Figure 1b) the PLS-DA revealed distinct separation of severe and critical patients, with 43.6% of the variance in the proportion of Mcs accounted for in the first component (Figure 2d). Mc25, 15, and 5 were enriched in critical infection, with Mc23 and 2 enriched in severe infection patients (Figure 2e & b). To confirm that the difference in these Mcs were not an artefact of the unsupervised nature of the PLS-DA, unpaired Mann-Whitney U-tests comparing the proportions of each Mc between severe and critical patients was performed. Mc25 (GZMB^+^PFN^+^ CD4^+^ CD45RO^-^, CCR7^-^), Mc15 (GZMB^+^PFN^+^ CD4^+^ CD45RO^+^, CCR7^-^), and Mc5 (HLA-DR^+^ CD4^+^ CD45RO^+^, CCR7^+^) were significantly enriched in critical compared to severe infection (Figure 2f). While the expression of CCR7 and CD45RO on Mc25 and 15 suggest a T_EMRA_ and T_EM_ phenotype respectively, these Mcs also appeared to express intermediate levels of CXCR5 on the heatmap, a feature of CD4^+^ T_FH_ cells (Figure 2c). However, Mcs 25 and 15 displayed a spectrum of expression of CXCR5 (Figure s2) and thus this analysis did not allow definitive determination of T_FH_ phenotype. Therefore, Mc25 and 15 represent cytotoxic CD4^+^ T lymphocyte subsets, with Mc5 being a population of activated T_CM_ cells. The proportion of Mc23 (CD8^+^ CD45RO^-^, CCR7^+^) and Mc2 (CD4^+^ CD45RO^-^, CCR7^+^) were significantly greater in severe patients than critical, representing non-activated naïve (T_N_) CD8^+^ and CD4^+^ T cells, respectively (Figure 2f). This analysis suggests an expansion of activated and cytotoxic CD4^+^ T cells populations and a decrease in the proportion of T_N_ cell subsets is involved in progression from severe to critical SARS-CoV-2 infection.

**Figure 2.**
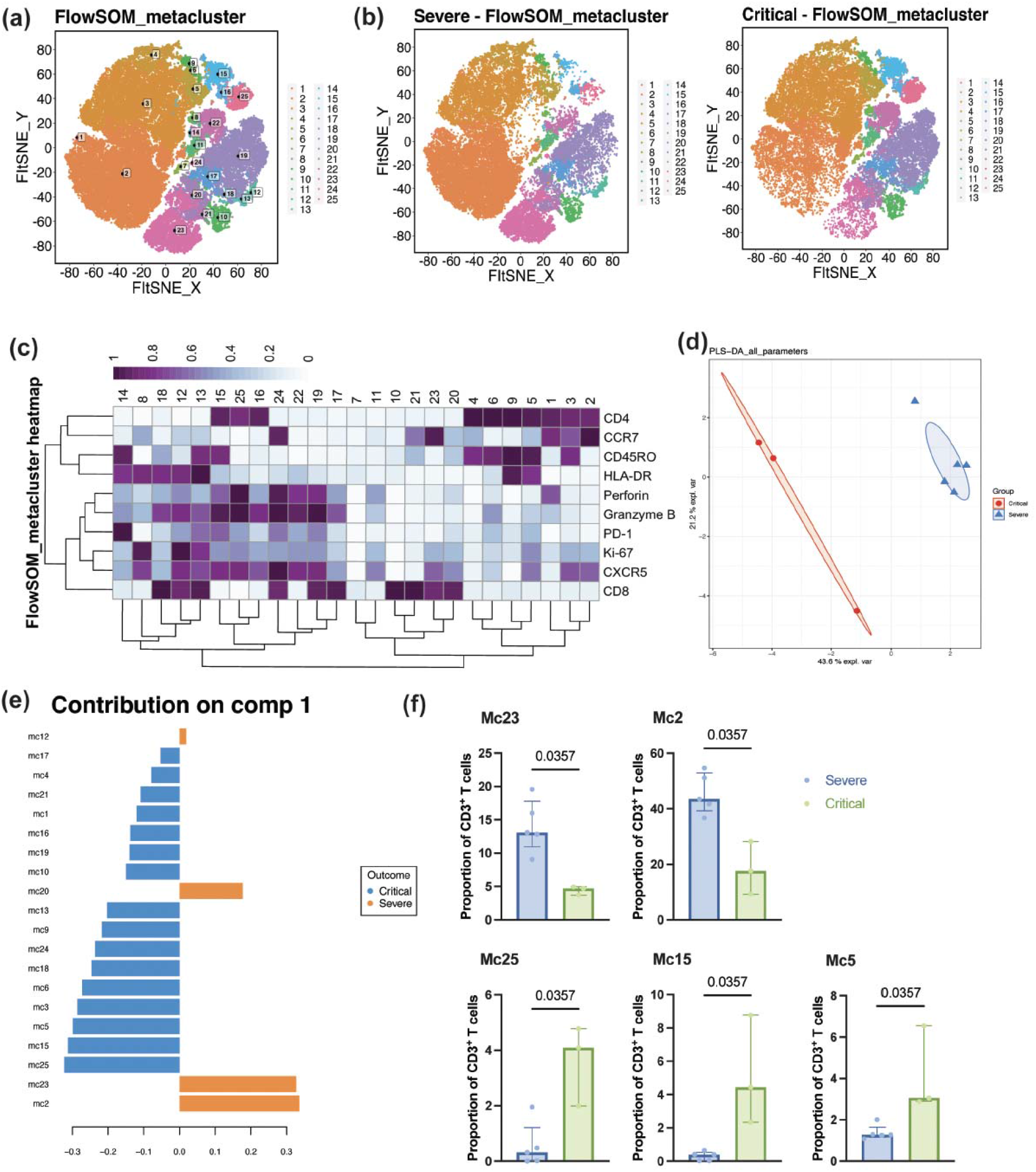
Unbiased clustering of T cell compartment in severe and critical patients. **A)** FIt-SNE visualisation of FlowSOM automatic clustering of a subsample of T cells from each severe (n = 5) and critical (n = 3) patient. **B)** FIt-SNE visualisation of alterations in proportion of metaclusters making up the T cell compartment between severe and critical disease. **C)** Heatmap plot showing the relative expression of each marker on self-organised map metaclusters. **D)** Partial Least Squares Discriminant Analysis of relative proportions of 20 non-redundant CD4^+^ and CD8^+^ T cell populations defined by automatic clustering. **E)** Christmas tree plot visualising the variables contributing to variance between severe and critical disease as determined by PLS-DA **F)** Non-parametric Mann-Whitney U test of proportions of metaclusters between severe and critical infection patients; error lines represent median ± interquartile range.

### Cytotoxic CD4^+^ T lymphocytes are characteristic of disease progression

As the proportions of naïve and memory T cells were negatively correlated (Figure 1c), the distribution of naïve/memory subsets were further explored in the CD4^+^ non-T_FH_ and CD8^+^ T cell compartments by manual gating on CD45RO and CCR7 (Figure 3a). Critical patients exhibited increased proportions of CD8^+^ T_EMRA_ cells and a reduction in the proportion of CD4^+^ T_N_ cells, compared to both mild and severe disease (Figure 3b). In contrast, the T cell compartments in asymptomatic, mild, moderate, and severe patients were composed of comparable proportions of T_N_ CD4^+^ and CD8^+^ cells. While the proportion of CD4^+^ T_EMRA_ cells was not significantly elevated, critical patients had increased expression of the activation marker HLA-DR on CD4^+^ T_EMRA_ cells compared to those with asymptomatic or mild disease (Figure 3c). There was also a notable increase in the proportion of CD8^+^ T_EMRA_ cells expressing the activation marker PD-1 as disease severity worsened (Figure 3c). As Mc25 and 15 could not clearly be defined as T_FH_ cell populations, manually gated CXCR5^hi^ CD4^+^ T_FH_ cells were selected for further analysis. While the proportion of total T_FH_ and CCR7^+^PD-1^-^ T_FH_ cells was not significantly different between groups, the proportion of circulating PD-1^+^CCR7^-^ T_FH_ cells (cT_FH_) was elevated in critical infection (Figure 3d).

**Figure 3.**
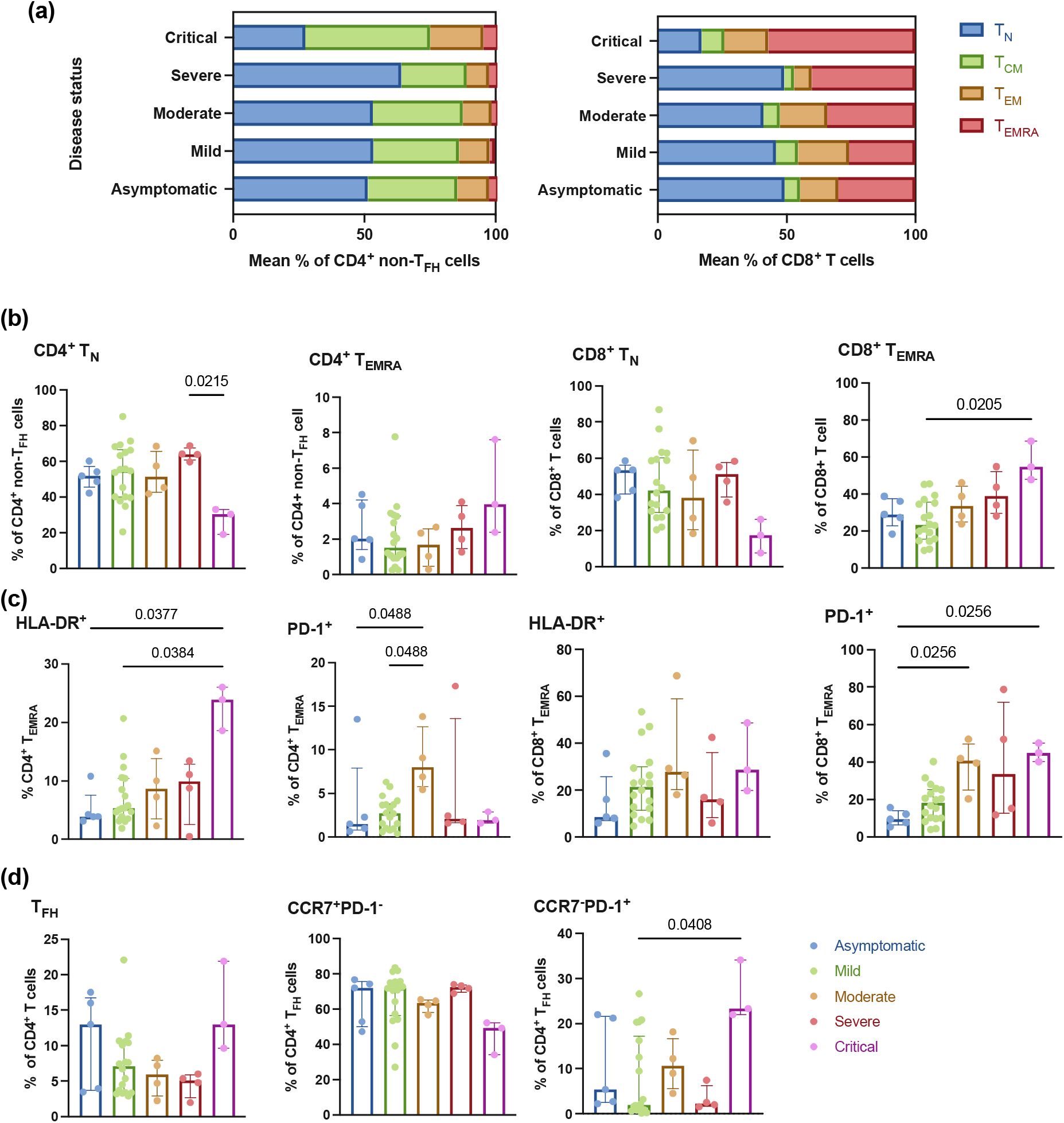
Differentiation status of CD4^+^ and CD8^+^ T cell compartments. **A)** Summary bar plots representing frequency of CD4^+^ and CD8^+^ T cell naïve and memory subsets between disease states defined as T_N_ (CCR7^+^CD45RO^-^), T_CM_ (CCR7^+^CD45RO^+^), T_EM_ (CCR7^-^CD45RO^+^), and T_EMRA_ (CCR7^-^CD45RO^-^). **B)** Frequency of CD4^+^ and CD8^+^ T_N_ and T_EMRA_ subsets **C)** Frequency of CD4^+^ and CD8^+^ T_EMRA_ cells expressing HLA-DR and PD-1. **D)** Frequency of total T_FH_ cells defined by CXCR5^hi^CD4^+^ phenotype, and CCR7^+^PD-1^-^ and CCR7^-^PD-1^+^ cT_FH_ cells. Difference between groups determined by non-parametric Kruskal-Wallis, with comparison of the rank mean of experimental groups by Original FDR method of Benjamini and Hochberg; error bars represent median ± interquartile range.

The computational analyses in Figures 1 and 2 revealed distinct populations of cytotoxic (GZMB^+^PFN^+^) CD4^+^ T cell subsets enriched in critical disease. As cytotoxic CD8^+^ T cells have been correlated with disease severity and mortality in COVID-19,^22^ the proportion of GZMB^+^PFN^+^ CD4^+^ and CD8^+^ subsets were of interest to investigate further. As a proportion of lymphocytes, critical patients had expanded populations of cytotoxic CD4^+^, but not CD8^+^ T cells. Cytotoxic CD4^+^ T cells made up a mean of 3.68% of the lymphocyte compartment in critical infection patients, and between 0.41-1.28% in all other disease states (Figure 4a). To delineate the subsets of cytotoxic CD4^+^ T cells contributing to this, the proportions of GZMB^+^PFN^+^ T_FH_ and CD4^+^ non-T_FH_ cells were compared across disease states. Both these subsets were significantly elevated in critical disease, and as such the phenotype of subpopulations within these subsets were explored further (Figure 4b). Of the CD4^+^ non-T_FH_ cell compartment, there was a higher proportion of CD4^+^ T_EMRA_ cells expressing GZMB^+^PFN^+^ cells in critical infection (Figure 4c). This was in line with the predominant T_EM_ and T_EMRA_ phenotype of cytotoxic compared to total CD4^+^ non-T_FH_ cells, which were largely of a T_N_ and T_CM_ phenotype (Figure 4d). Similarly, cytotoxic CD4^+^ T_FH_ cells were almost exclusively of a CCR7^-^PD-1^+^ cT_FH_ phenotype and total T_FH_ cells were predominantly of a CCR7^+^PD-1^-^ phenotype (Figure 4e). Finally, the proportion of cytotoxic CD4^+^ and CD8^+^ T cells was correlated to determine whether CD4^+^ cytotoxicity may be a compensating for CD8^+^ T cell exhaustion. However, these two populations correlated positively with each other (p = 0.0039: Figure 4f). From these data it appears that the expansion of cytotoxic CD4^+^ cT_FH_ and T_EMRA_ populations during SARS-CoV-2 infection is unique to critical disease.

**Figure 4.**
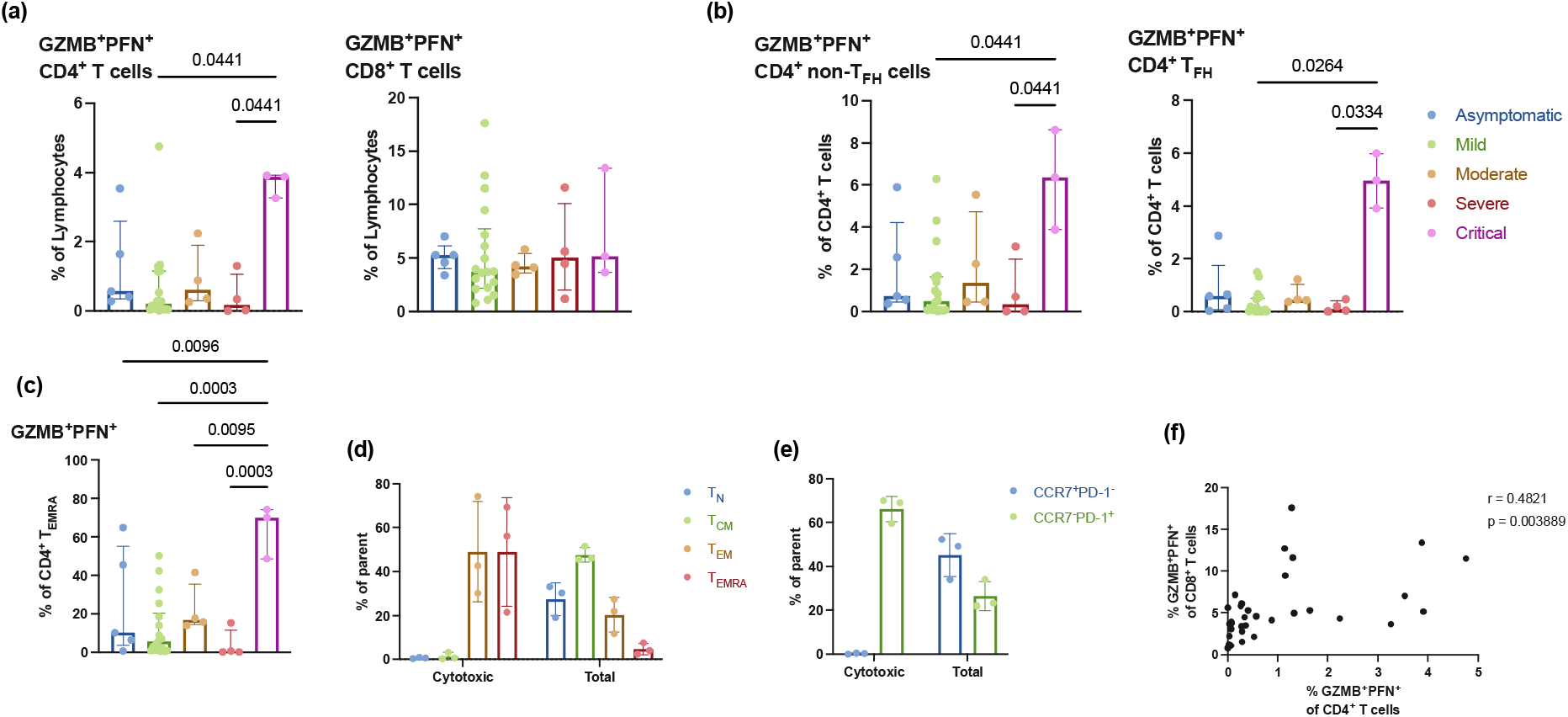
Cytotoxic CD4^+^ and CD8^+^ T cells are expanded in critical infection. A) Frequency of GZMB^+^PFN^+^ CD4^+^ and CD8^+^ T cells across disease states. B) Frequency of GZMB^+^PFN^+^ non-T_FH_ cells (CXCR5^-/lo^) and T_FH_ cells (CXCR5^hi^) as a proportion of all CD4^+^ T cells C) Frequency of GZMB^+^PFN^+^ CD4^+^ T_EMRA_ cells as a proportion of parent D) Summary bar plot representing the differentiation status of total CD4^+^ non-T_FH_ cells and cytotoxic (GZMB^+^PFN^+^) CD4^+^ T cells defined as T_N_ (CCR7^+^CD45RO^-^), T_CM_ (CCR7^+^CD45RO^+^), T_EM_ (CCR7^-^CD45RO^+^), and T_EMRA_ (CCR7^-^CD45RO^-^) in critical infection patients (n = 3). E) Summary bar plot representing the differentiation status of total CD4^+^ T_FH_ (CXCR5^+^) cells and cytotoxic (GZMB^+^PFN^+^) CD4^+^ T_FH_ cells by CCR7^+^PD-1^-^ and CCR7^-^PD-1^+^ (cT_FH_) cell phenotype in critical infection patients (n = 3). F) Pearson correlation between GZMB^+^PFN^+^ CD4^+^ and CD8^+^ T cells as a proportion of total CD4^+^ and CD8 T^+^ cells, respectively. Difference between groups determined by non-parametric Kruskal-Wallis, with comparison of the rank mean of experimental groups by Original FDR method of Benjamini and Hochberg; error bars represent median ± interquartile range.

## Discussion

Characterising immune responses that associate with different disease severities of COVID-19 may help to define protective immune responses to SARS-CoV-2 and guide the rational development of next-generation vaccines. This study provides a phenotypic analysis of the T cell response in patients experiencing asymptomatic to critical SARS-CoV-2 infection. We confirm that the T cell compartment is distinctly altered in critical SARS-CoV-2 infection, defined by an expansion of effector memory subsets, and increased expression of activation and cytotoxic functional markers on CD4^+^ T cells. These data suggest a potentially pathogenic role of cytotoxic CD4^+^ CTLs in the progression of COVID-19.

CD4^+^ T cells are well-established critical responders in viral infection; however, CTL function is more commonly associated with CD8^+^ T cell populations. As such, it is of interest that the top Mcs associated with the progression of disease from severe to critical were activated and cytotoxic CD4^+^ T cell populations (Figure 2). CD4^+^ T cells can mediate host-cell death through secretion of granzyme B and perforin^24,25^, and CD4^+^ CTLs have been identified during viral infections including human immunodeficiency virus (HIV), human cytomegalovirus (CMV), Epstein Barr virus (EBV), influenza, dengue virus and more recently SARS-CoV-2.^26^ In patients with COVID-19, CD4^+^ T cells expressing high levels of *PFN1, GZMB* and *GZMH* transcripts have been identified previously by scRNA-seq to be enriched in hospitalised compared to non-hospitalised COVID-19 patients.^27^ The data provided in this current study describes an increase in CD4^+^ CTLs, and a unique expansion of GZMB^+^PFN^+^ CD4 T_EMRA_ cells during critical infection at the level of protein expression. While the circulating T cell populations have been analysed in this cohort, high infiltration of CD4^+^ CTLs, as well as CD8^+^ CTLs, have also been reported in the lung parenchyma of severely ill COVID-19 patients.^24,28^ The elevated expression of HLA-II in the respiratory epithelium provides a potential mechanistic basis for the role of CD4^+^ CTLs in the profound host tissue damage associated with SARS-CoV-2 acute respiratory distress syndrome (ARDS).^24^

In the CD4 T_FH_ cell compartment, critical patients also exhibited increased proportions of cytotoxic cells, which were predominantly of a CCR7^-^PD-1^+^ phenotype; such cells have been previously described as circulating T_FH_ (cT_FH_) cells.^29^ Cytotoxic T_FH_ cells have been shown to induce B cell death and to correlate negatively with antibody titres in recurrent Strep A infection in children.^30^ Post-mortem investigations have shown loss of germinal centre (GC) B cells and absence of GCs in the lymph nodes of COVID-19 decedents. ^31^ However, while cytotoxic cT_FH_ cells were elevated and correlated negatively to anti-S1/S2 SARS-CoV-2 antibodies in hospitalised patients, this correlation was not seen in non-hospitalised patients.^27^ Conversely, antibody-secreting plasmablasts have been correlated with mortality in COVID-19,^22^ and higher antibody responses are associated with more severe disease.^32^ As there are limited studies investigating cytotoxic cT_FH_ in COVID-19, the implications of cytotoxicity in cT_FH_ cells require further investigation to assess any potential detrimental effect this cell type may have on antibody responses to SARS-CoV-2.

CD4^+^ T cell cytotoxicity has been proposed to be a compensatory mechanism to combat exhaustion of CD8^+^ T cells, in which CD8^+^ CTL expression of GZMB and PFN decrease, and PD-1 increases.^33,34^ In contrast, no change in the proportion of CD8^+^ GZMB^+^PFN^+^ subsets was observed here across disease severities. Moreover, PD-1 expression was not highlighted as a key variable in the separation of disease states in the PCA. This suggests that if CD8^+^ CTL exhaustion was present it was not a differentiating feature of disease progression. As such, the increased proportion of PD-1^+^CD8^+^ T_EMRA_ cells in critical infection likely represents a phenotype of increased activation, rather than functional exhaustion. In line with this view, previous studies have reported elevated frequencies of hyperactivated CD8^+^ cells, defined by expression of HLA-DR^+^CD38^+^PD-1^+^TIM-3^+^, in severe and critical SARS-CoV-2 infection.^21,22^ While no difference in cytotoxicity was reported here, GZMB and PFN expression has been correlated with critical disease and mortality in COVID-19.^22,35–39^ Rha et al., (2021) reported GZMB and PFN were expressed by almost all SARS-CoV-2-specific multimer^+^ CD8^+^ T cells.^40^ As such, enrichment for SARS-CoV-2 specific T cells may provide more insight into the changes in proportion and phenotype of cytotoxic CD8^+^ T cells between disease states. In combination with these studies, the data presented here suggest that hyperactivated effector memory subsets of CD8^+^ T cells may contribute to COVID-19 progression.

Investigating the immune response in asymptomatic and mild disease can shed light on the immune response that effectively controls viral replication and disease progression. Control of viral load by T cells was demonstrated in B-cell depleted Rhesus Macaques ^41^, and CD8^+^ T cell responses correlated with better clinical outcome in patients with inborn errors in humoral immune responses and B cell impairment in SARS-CoV-2 infection.^42,43^ In the current study, asymptomatic and mild patients exhibited low frequencies of activated or cytotoxic cells, and a predominantly naïve phenotype in both the CD4^+^ and CD8^+^ compartments. However, there is an inherent limitation to investigating the peripheral immune response in a respiratory infection where localised inflammation and resident immune cell responses may not be reflected in circulation.^44^ Furthermore, this study did not investigate SARS-CoV-2-specific T cells, and as such a proportionally smaller T cell response may be missed in these less severe disease states. It has been shown previously that individuals with mild infection are less likely to have detectable SARS-CoV-2-specific T cell responses.^45^ In combination with the relative lack of a robust T cell response in asymptomatic and mild disease here, this may question the necessity of a strong T cell response to prevent the progression of COVID-19. This is consistent with the diverse polyclonal, less differentiated SARS-CoV-2 specific T cell response observed in children with mild and asymptomatic T cell response than the clonally expanded memory T cell response with markers of cytotoxicity and exhaustion present in adults.^46^ The protective capacity of T cells should be investigated further to assess whether preserved T cell response against variant SARS-CoV-2 viruses provide protection against severe disease, as has been suggested previously. ^12,13,16^

Finally, there are several clinical variables that may confound this analysis, notably the impact of age on the immune response to infection. The critical patients were older than asymptomatic and mild patients which is reflective of the high median age of COVID-19 patients admitted to hospital and ICU.^47^ Age over 65 years is associated with a progressive decline in immune function characterised by decreased thymic function, contraction of naive T cell populations, and perturbed T cell function, such as reduced cytokine production and proliferative capacity.^48,49^ It has been shown previously that SARS-CoV-2 immune dysregulation mimics that seen in age-related immunosenescence.^50^ Without age-matched patients, the impact of age cannot be extricated from that of disease severity on the T cell response to SARS-CoV-2.

The data presented in our study add to our understanding of the contribution of T cell responses to disease progression in SARS-CoV-2 infection. The distinct absence of a notable T cell response in asymptomatic disease but expansion of cytotoxic CD4^+^ cT_FH_ and T_EMRA_ subset in critical disease, suggest that CTLs may be contributing to host tissue damage and systemic inflammatory disease. As such, the potentially detrimental role of T cell responses in COVID-19 should be considered in the development of next-generation therapies and vaccines against SARS-CoV-2.

## Material and Methods

### Patient Demographics

SARS-CoV-2 PCR-positive patients and their household contacts were enrolled through the Royal Prince Alfred (RPA) hospital COVID-19 clinic or virtual care system in March of 2020 (COVIMM cohort). Ethics approval was granted by the RPA ethics committee, human ethics number X20-0117 and 2020/ETH00770. Verbal consent was given by all participants. Additional patients enrolled in the COSIN study in June 2021 through seven hospital microbiology labs, and out-patient care units across Sydney, Australia, as described by Balachandran et al. (2022) were also included. Ethics approval was granted by Human Research Ethics Committees of the Northern Sydney Local Health District and the University of New South Wales, NSW Australia (ETH00520), and written consent was obtained from all patients.

SARS-CoV-2 infection was defined by a positive nasopharyngeal RT-PCR performed by accredited laboratories within the Sydney Health District. Patients were classified as asymptomatic (n = 5), mild (n = 18), moderate (n = 4), severe (n = 5), or critical (n = 3) (Supplementary Table 1). COVID-19 disease severity as defined by the NIH guidelines (https://www.covid19treatmentguidelines.nih.gov/overview/clinical-spectrum/). Patients were classified as mild if their symptoms were self-manageable.

### Immunophenotyping by spectral cytometry

Patient PBMCs were isolated from whole blood by Ficoll-density gradient separation, and cryopreserved in Heat-inactivated FBS with 10% DMSO at -80°c. Cryopreserved PMBC samples were thawed and diluted to a concentration of 1 × 10^6^ live PMBCs in RMPI supplemented with 10% FCS and 100 ug/ml streptomycin and penicillin in a U-bottom 96-well plate. Cells were washed in FACS buffer, dead cells labelled with LIVE/DEAD™ Fixable Blue dye (Invitrogen) for 20 minutes at 4°C and washed twice in FACS buffer. Cells were stained with extracellular markers (Supplementary Table 2; CD3-BUV395 (1:20), CD4-PerCPCy5.5 (1:100), CD8-BUV496 (1:100), CD19-BV510 (1:50), CD56-BV605 (1:50), HLA-DR-BV650 (1:50), CD45RO-BV786 (1:50), CD197-PECy7 (1:20), and CXCR5-AF647 (1:20) in FACS buffer for 20 minutes at 4°C. Cells were then permeabilised and fixed with Transcription Factor Buffer Set (BD Pharmingen™) for 40 minutes at 4°C. Cells were washed then stained with intracellular markers (Supplementary Table 2) PD-1-BV737 (1:50), Ki-67-BV711 (1:50), Perforin-PE (1:50), and Granzyme B-APC (1:20) in Transcription Factor wash buffer (BD Pharmingen™). Anti-Mouse Ig, κ/Negative Control Particles Set (BD™ CompBeads) were used for single stain controls and stained with corresponding extracellular and intracellular stains. Fluorescent minus one (FMO) controls were included for intracellular markers and stained at the concentration of patient samples at corresponding staining times. The samples underwent a second fixation step with 4% paraformaldehyde for 30 minutes, washed, and resuspended in 200 ml FACS buffer for acquisition. Patient samples and controls were acquired using the 5-laser Cytek Aurora® Spectral Cytometer. Spectral unmixing was performed using the inbuilt SpectroFlo® software after acquisition of unstained and single-stained controls and before patient sample acquisition.

### Data Analysis

Data analysis was performed in FlowJo v10.8.1, R v4.2.1, and GraphPad Prism v9. Dead cells, doublets, and debris were excluded in FlowJo v10.8.1 by FSC-A, FSC-H, SCC-A, and LIVE/DEAD dye (Figure S1). NKT and B cells were excluded by CD3^+^CD56^+^ and CD3^-^CD19^+^ phenotype, respectively. Compensation for unmixing errors was performed in FlowJo v10.8.1. Irregularities in staining and acquisition between batches were controlled by matching gates on control samples and applying identical gate lineage and functional marker expression within each staining batch.

To control for the differences between staining batches, the Principal Component Analysis (Figure 1) was calculated using 38 overlapping CD3^+^ T cell populations that were manually gated in FlowJo v10.8.1. CD4^+^ and CD8^+^ T cells were first divided into T_N_ (CCR7^+^CD45RO^-^), T_CM_ (CCR7^+^CD45RO^+^), T_EM_ (CCR7^-^CD45RO^+^), T_EMRA_ (CCR7^-^CD45RO^-^) subsets. The frequency of CD4^+^ and CD8^+^ T_N_, T_CM_, T_EM_, and T_EMRA_ subsets, and the cells expressing HLA-DR, PD-1, GRZMB, PFN, and Ki-67 in the T_CM_, T_EM_, and T_EMRA_ subsets were included to assess the significance of T cell function on disease status (Figure S2). Each population was calculated as a proportion of its respective CD4^+^ or CD8^+^ T cell compartment, and these frequencies were used for PCA analysis in R v4.2.1 using the Spectre package.^51^

For the comparison between severe and critical infection, the R package Spectre was utilised for computational analyses (https://www.ncbi.nlm.nih.gov/pubmed/33840138). CD3^+^ T cells were exported for each patient sample. FlowSOM clustering created 25 metaclusters. Fit-SNE dimensionality reduction^52^ was run on subsampled data of 50,000 cells from severe and critical groups to create a representative plot between disease states and a heatmap plot of cellular marker expression on each metacluster was created using the pheatmap function. To identify contributions to differences between severe and critical infection, a partial least squares discriminant analysis (PLS-DA) was done.^53^ To validate the phenotype of metaclusters, an FCS file of marker expression on each metacluster was created using the write.files function in Spectre. This allowed for clear analysis of marker expression for each metacluster in FlowJo v10.8. After exclusion of CD4^-^CD8^-^ FlowSOM metaclusters, PLS-DA was performed to identify the variability in proportion of metaclusters between severe and critical disease. Validation of the statistical differences in proportion of metaclusters between groups was performed in GraphPrism v9.

The difference between groups were analysed by two-sided paired Mann-Whitney U-test, and non-parametric Kruskal-Wallis, with comparison of the rank mean of experimental groups by Original False Discovery Rate (FDR) method of Benjamini and Hochberg. Statistical significance between groups in computational analyses were calculated with a permutation ANOVA, with correction for multiple comparisons by FDR method. Statistical significance was set as p ≤ 0.05.

## Data Availability

The datasets generated during and/or analysed during the current study are available from the corresponding author on reasonable request.

## Acknowledgments

This work was supported by MRFF COVID-19 Vaccine Candidate Research Grant 2007221 (J.A.T., C.C., M.S., A.L.F.). F.M._W. and T.M.A. are supported by the International Society for the Advancement of Cytometry (ISAC) Marylou Ingram Scholars program. COSIN cohort was supported by Snow Medical Foundation as an investigator-initiated study. We acknowledge the support of the University of Sydney Advanced Cytometry Facility. The authors thank the study participants for their contribution to this research and the clinical staff who collected the samples.

## Figure Legends

**Supplementary Figure 1.**
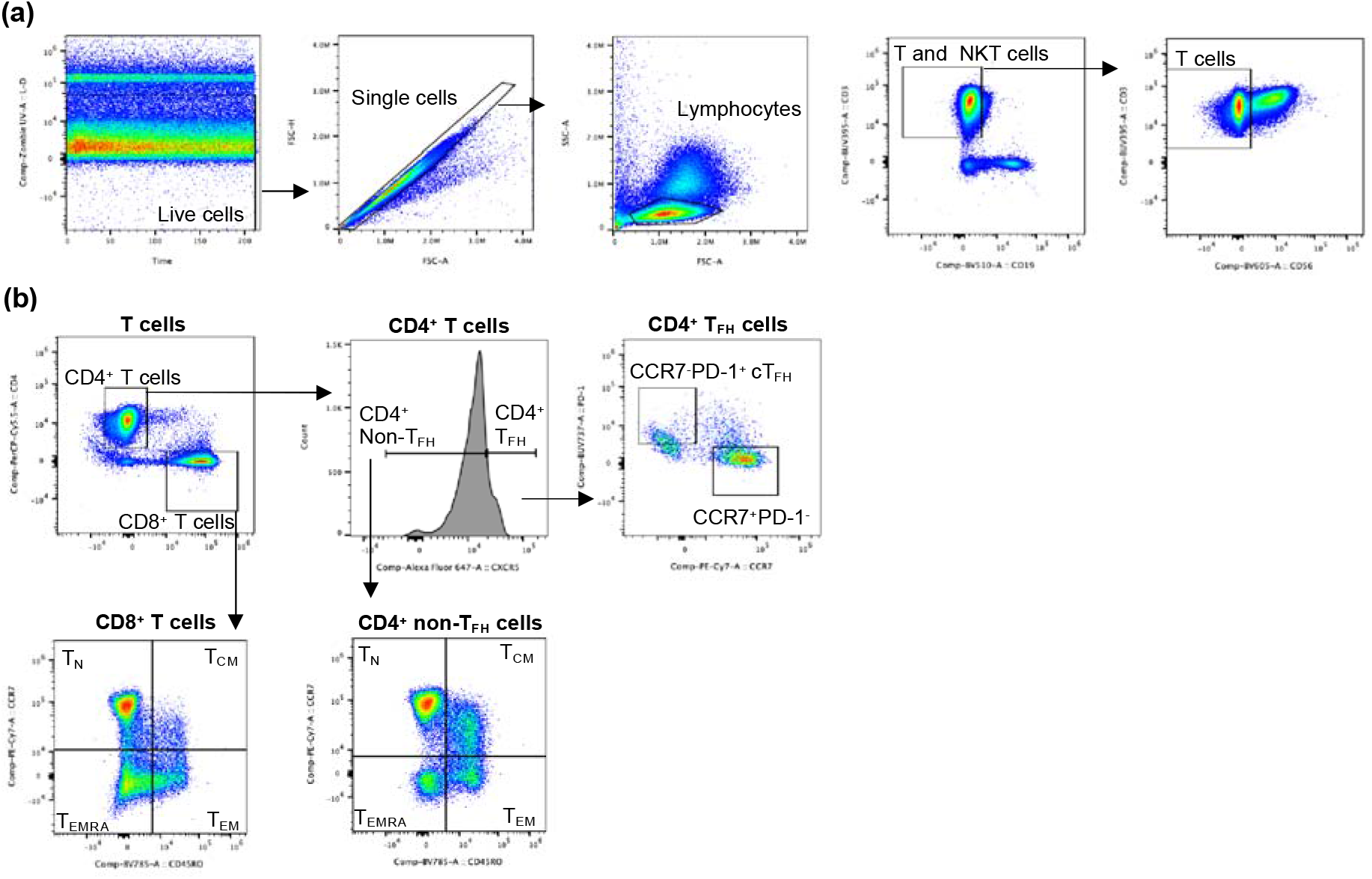
Gating strategy for T cells subsets. Human peripheral blood mononuclear cells (PBMCs) were analysed by spectral cytometry. **A)** Dead cells, cell aggregates, and myeloid cells were excluded by LIVE/DEAD, time, FSC-H, FSC-A, and SSC-A. T cells were isolated by CD3^+^CD19^-^ and CD3^+^CD56^-^ phenotype to exclude B cells and NKT cells, respectively. **B)** T cells were divided into CD4^+^ and CD8^+^ T cells. CD4^+^ T cells were divided further into CXCR5^-/lo^ CD4^+^ non-T_FH_ cells and CXCR5^hi^ T_FH_ cells. T_FH_ cells were defined as cT_FH_ by CCR7^-^ PD-1^+^ and as CCR7^+^PD-1^-^ cells. CD4^+^ non-T_FH_ and CD8^+^ T cells were divided into naïve/memory subsets defined as T_N_ (CCR7^+^CD45RO^-^), T_CM_ (CCR7^+^CD45RO^+^), T_EM_ (CCR7^-^CD45RO^+^), and T_EMRA_ (CCR7^-^CD45RO^-^). Flow plots of representative non-infected control sample COVIMM_093 and performed in FlowJo v10.8.1.

**Supplementary Figure 2.**
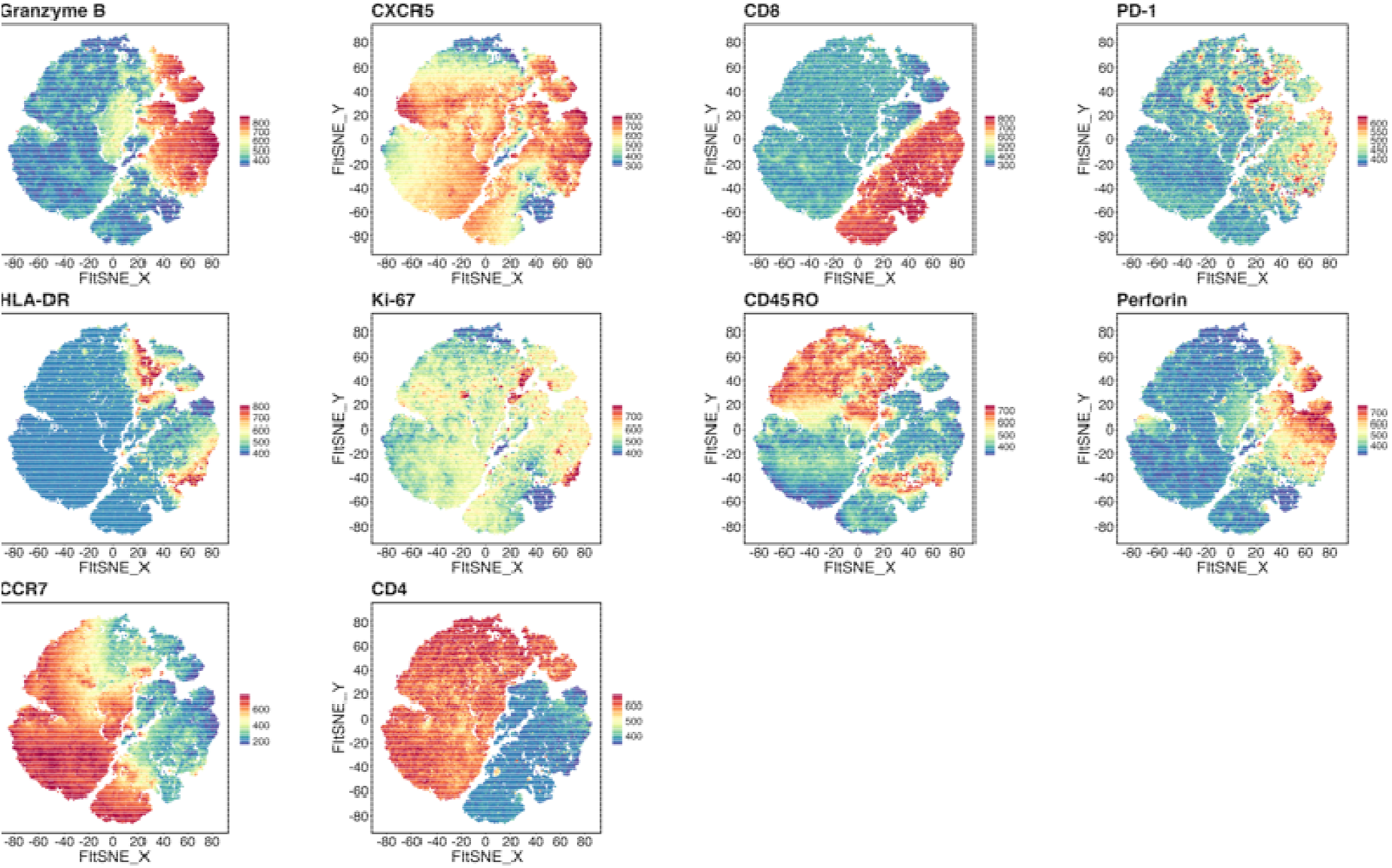
Expression of lineage and functional markers on FIt-SNE of metaclusters. Relative expression of cellular marker expression on FIt-SNE visualisation of FlowSOM automatic clustering of a subsample of T cells from each severe (n = 5) and critical (n = 3) patient.

**Supplementary Table 1.** Patient clinical characteristics

| **Peak disease status (n)** | **Median age**<br>**(range)** | **Days post**<br>**positive swab (range)** |
| --- | --- | --- |
| Asymptomatic (5) | 30 (25-51) | 11 (5-32) |
| Mild (18) | 31 (12-60) | 15 (5-49) |
| Moderate (4) | 58 (55-79) | 21 (14-28) |
| Severe (5) | 49 (24-69) | 14 (14-112) |
| critical (3) | 65 (59-65) | 28 (28) |
| **Total** | **35 (12-79)** | **14 (5-12)** |

**Supplementary Table 2.** Antibody Panel ^†^Markers used for automatic clustering; ^‡^ Intracellular markers

| Product | Fluorophore | Clone | Detector | Species raised in | Source |
| --- | --- | --- | --- | --- | --- |
| CD3 | BUV395 | SK7 | UV375_C | Mouse | BD Horizon™ |
| Invitrogen LIVE/DEAD™ Fixable Blue Dead Cell Stain Kit, for UV excitation | N/A | N/A | UV450 | Mouse | Invitrogen |
| CD8<sup>†</sup> | BUV496 | RPA-T8 | UV514 | Mouse | BD Horizon™ |
| PD-1<sup>††</sup> | BUV737 | EH12.2 | UV737_ | Mouse | BD Horizon™ |
| CD19 | BV510 | HIB19 | V510_ | Mouse | BioLegend® |
| CD56 | BV605 | NCAM16.2 | V598 | Mouse | BD Biosciences® |
| HLA-DR<sup>††</sup> | BV650 | G46-6 | V660_C | Mouse | BD Biosciences® |
| Ki-67<sup>††</sup> | BV711 | Ki67 | V720 | Mouse | BioLegend® |
| CD45RO<sup>†</sup> | BV786 | UCHL1 | V780_A | Mouse | BioLegend® |
| CD4<sup>†</sup> | PerCPCy5.5 | RPA-T4 | B695_A | Mouse | BioLegend® |
| Perforin<sup>††</sup> | PE | B-D48 | YG582_D | Mouse | BioLegend® |
| CD197(CCR7)<sup>†</sup> | PECy7 | G043H7 | YG780_A | Mouse | BioLegend® |
| Granzyme B<sup>††</sup> | APC | QA16A02 | R660_C | Mouse | BioLegend® |
| CXCR5<sup>†</sup> | AF647 | RF8B2 | R679 | Rat | BD Biosciences® |

